# Biosecurity uptake and perceived risk of avian influenza among people in contact with birds

**DOI:** 10.1101/2025.04.23.25326059

**Authors:** Harry Whitlow, Suzanne Gokool, Genevieve Clapp, Irene Bueno, Mariam Logunleko, Peter Moore, Sarah Masterton, Jo Taylor-Egbeyemi, Ian Brown, Riinu Pae, Louise E Smith, Ellen Brooks-Pollock, Amy C Thomas

## Abstract

**Introduction:** Recent intercontinental spread of highly pathogenic avian influenza (HPAI) A(H5N1) among kept and wild birds, and transmission to mammalian hosts, including cattle and humans, has heightened the need to review public health risk assessments. Biosecurity measures (BMs) are essential for limiting disease spread, but how widely different practices are implemented is not fully known.

**Methods:** Here, we report on the uptake of BMs and risk perception of avian influenza virus (AIV) in the context of preventing zoonotic transmission to persons potentially at high risk of exposure. Questionnaire data from 225 people in contact with birds in the UK (Avian Contact Study, May to July 2024) were analysed.

**Results:** We found hand washing after contact with birds was the most common BM implemented (89%, 196 of 218), followed by using disinfecting footwear dips (78%, 170 of 218). Individuals in contact with a higher number of birds were more likely to use at least one personal protective equipment (PPE) measure for the face or body or at least one footwear- related PPE measure. Perceived risk of AIV to bird health was high for individuals in contact with large flocks (≥1001 birds) and associated with uptake of at least one footwear-related PPE measure (independent of flock size). Perceived risk of AIV to respondents’ own health was low, regardless of the number of birds a respondent had daily contact with.

**Conclusions:** Our results indicate that routinely used BMs are implemented to limit AIV spread among birds, but not with the purpose of limiting zoonotic transmission from birds to humans. Identifying cohort characteristics which could lead to low BM uptake, alongside barriers and facilitators to BM uptake is important for informing zoonotic AIV public health campaigns.

## Introduction

Between 2021 and 2022, 48 million birds were culled across Europe following outbreaks of highly pathogenic avian influenza (HPAI) A(H5N1) virus, belonging to clade 2.3.4.4b, in commercial poultry establishments and wild bird populations (1). Rapid and widespread A(H5N1) transmission in Europe (1), Africa (2), North America (3), South America and the Antarctic (4,5), constituted a panzootic in wild birds (3). In the UK, a total of 9.38 million birds were estimated to have been culled in June-December 2022 (6). To minimise avian transmission, in October 2022 the English Department for Environment, Food and Rural Affairs (Defra) and the Animal and Plant Health Agency (APHA) introduced a mandatory avian influenza prevention zone (AIPZ) and on-farm biosecurity measures across England (7). Transmission among domestic poultry (kept birds) subsided during 2024 (8), but resurgence as of January 2025 has led to the declaration of another AIPZ in England, demonstrating the virus’ persistent threat to domestic and wild animals (9).

Risk of avian influenza virus (AIV) infection for the general population in the UK remains low (10), but concern and risk assessments are heightened given A(H5N1) infection of wild and marine mammals (4), US dairy cattle (11), and higher mortality rates among wild birds (12). In humans, infection with A(H5N1) is rare following exposure to infected animals and no evidence of human-to-human transmission has been recorded (13). In 2023, an asymptomatic surveillance study detected four UK A(H5N1) cases in people exposed to AIV infected birds, although two were suspected to be due to contamination (10). One further case was detected in January 2025 (14). In the US, A(H5N1) has infected at least 1,077 dairy herds across 17 states with known spillover to 70 people (11,15); human infection has been either asymptomatic or mild, with one fatality in an individual ≥65 years with underlying health conditions (last updated 1 July 2025) (16). Despite the current low public health risk, genetic evolution and/or co-infection with human seasonal influenza virus increases the risk of virus adaption to humans (17). As it is difficult to limit transmission among wild birds and mammals, biosecurity measures that prevent the introduction of AIV to kept bird populations, and subsequently from those birds to humans, remain crucial for preventing AIV transmission.

Biosecurity measures (BMs) for individuals in contact with birds are recommended to prevent AIV transmission in multiple settings (18–20). A recent qualitative study in the UK found high uptake of footwear dips among commercial poultry producers (21), contrasting with backyard flock keepers who largely report non-compliance with Defra enforced BMs, including footwear dips (22). This demonstrates diverse uptake trends among different demographics in contact with birds, requiring assessment of all possible groups to inform a more nuanced understanding of biosecurity. Currently, BM uptake is largely focused on the prevention of on-farm avian transmission (23,24), resulting in a limited understanding of BM uptake to prevent avian-to- human transmission.

BM uptake among people in contact with birds can be influenced by the perceived risk of zoonotic disease transmission and AIV in birds (25). Protection motivation theory (26) suggests that uptake of BMs is driven by perceived susceptibility and severity of AIV infection in birds, alongside the effectiveness of actions intended to prevent transmission (response efficacy). Studies in Nigeria and China have shown that biosecurity uptake to prevent avian-to-avian transmission varies by education status, occupation duration and farming operation size (23,27). In Taiwan and Europe, poultry farmers have documented personal attitudes, knowledge and concern for business viability to influence uptake of BMs (24,28,29). Factors such as age, gender and education have also been shown to either influence or have no effect upon AIV awareness and knowledge for both avian- and human-specific BMs in Europe, Vietnam and China (24,30,31). Moreover, existing studies on motivation are concentrated in South East Asia (31,32), limiting generalisability to the UK. This suggests that the uptake and source of motivation for BMs is heterogenous and requires investigation regarding avian-to-human transmission within a UK context.

Using data from the Avian Contact Study, a cross-sectional questionnaire of individuals in contact with kept and wild birds in the UK (33), we investigate trends in BM uptake and risk perception of AIV in the context of preventing zoonotic influenza infection. Building upon previous research regarding farming operation size (31) and the influence of risk perception upon BM uptake (25), we hypothesised that the number of birds a person had contact with on a typical day (‘bird contact’) and the perceived risk of AIV may influence uptake of different types of measures to prevent human infection with AIV.

## Materials and methods

### Brief description of the Avian Contact Study

The Avian Contact Study (www.bristol.ac.uk/avian-contact) (33) aimed to inform current zoonotic influenza public health measures given increased HPAI A(H5N1) transmission in birds. A cross-sectional, self-reported questionnaire was co-developed with avian influenza public health teams (UK Health Security Agency; UKHSA), veterinary government scientists (Animal and Plant Health Agency; APHA) and poultry farmers. This questionnaire was designed to capture the knowledge and practices of people who have contact with kept and/or wild birds in the UK, and therefore responses could have been given in reference to either. Eligible participants were aged 18 years or over, a UK resident (England, Northern Ireland, Scotland, Wales) and had contact with domestic and/or wild birds. Full details of the questionnaire methodology can be found in the Data Note (33), and the questionnaire is available as extended data (34).

### Ethics and consent

Ethical approval for the study was obtained from the University of Bristol, Faculty Research Ethics Committee, approval number 17048 on 16 January 2024.

Informed written consent (using e-consent hosted on Research Electronic Data Capture (REDCap) tools (35,36)), for the use of data collected via the questionnaire was obtained from respondents.

### Data collection

We used a convenience sampling method, where the questionnaire was initially delivered in- person at the British Pig and Poultry Fair (15 and 16 May 2024) - an agricultural conference aimed at pig and poultry keepers in the UK (37). The online questionnaire was also distributed via individuals met at the fair, animal health protection teams from APHA and poultry networks on social media. All questions were optional. Questionnaire data were collected using the Research Electronic Data Capture (REDCap) platform, hosted at the University of Bristol (35,36). Data captured between 15 May 2024 to 31 July 2024 were included in these analyses.

The questionnaire captured demographic data including age, gender, occupation and health. Occupational titles were derived from the Office for National Statistics’ Extended Standard Occupational Classification 2020 Framework (38). Bird contact and ownership questions presented categories of increasing frequencies of birds, BM uptake questions gave checkboxes to produce a binary outcomes and risk perception questions showed a likert scale of increasing risk perception options. We analysed the following questions relating to bird contact, biosecurity measures and risk perception:

- **How many birds of any type were you in direct contact with on a typical day?**

- 1-10
- 11-100
- 101-1000
- 1001-10,000
- 10,001-100,000
- More than 100,000
- **If you are a bird owner, how many birds do you own?**

- 1-10
- 11-100
- 101-1000
- 1001-10,000
- 10,001-100,000
- More than 100,000+
- I am not a bird owner
- **Do you use any biosecurity measures to limit the risk of catching avian influenza (bird flu)? Please select all that apply.**

- I use face masks
- I use goggles
- I use gloves
- I change clothing when in contact with birds
- I use outer garments when in contact with birds
- I change boots
- I use boot covers
- I use a disinfecting footwear dip
- I wash/clean my hands after touching birds
- I wash/clean my hands before and after handling raw poultry meat
- I make sure raw poultry meat is fully cooked before eating
- I do not eat undercooked or raw poultry meat
- I make sure eggs are thoroughly cooked
- I do not eat raw eggs
- I do not remember
- None
- Other
- **How much risk do you think avian influenza (bird flu) currently poses to your physical health?**

- Very high risk
- High risk
- Medium risk
- Low risk
- No risk at all
- I do not know
- **How much risk do you think avian influenza (bird flu) currently poses to the health of your birds?**

- Very high risk
- High risk
- Medium risk
- Low risk
- No risk at all
- I do not know

### Data analysis

Data were pre-coded in the REDCap software and exported to R Studio (R version 4.1.1) for data cleaning and analyses. Pre-processing steps involved assigning factor levels and labels for categorical variables. When an appropriate job title was not listed, respondents could input their own job title as free-text – this was manually cleaned and consolidated into existing appropriate occupational categories. Occupations with fewer than five individuals per category are not shown to preserve anonymity. Due to low counts among categories of bird contact, BM uptake and risk perception of AIV responses were categorised according to the groupings shown in Table 1. Respondents who selected “I do not know” for the questions relating to risk perception of AIV to physical health and bird health were considered as missing and excluded from subsequent analyses. The frequency distribution of BM uptake and risk perception variables before grouping is shown in the supplementary information (Figures S1 and S2). Use of BMs were recorded using checkboxes that respondents ‘checked’ for yes or left blank for ‘no’. We assumed responses with all measures ‘unchecked’ indicated a skipped question and were therefore coded as missing.

**Table 1.**
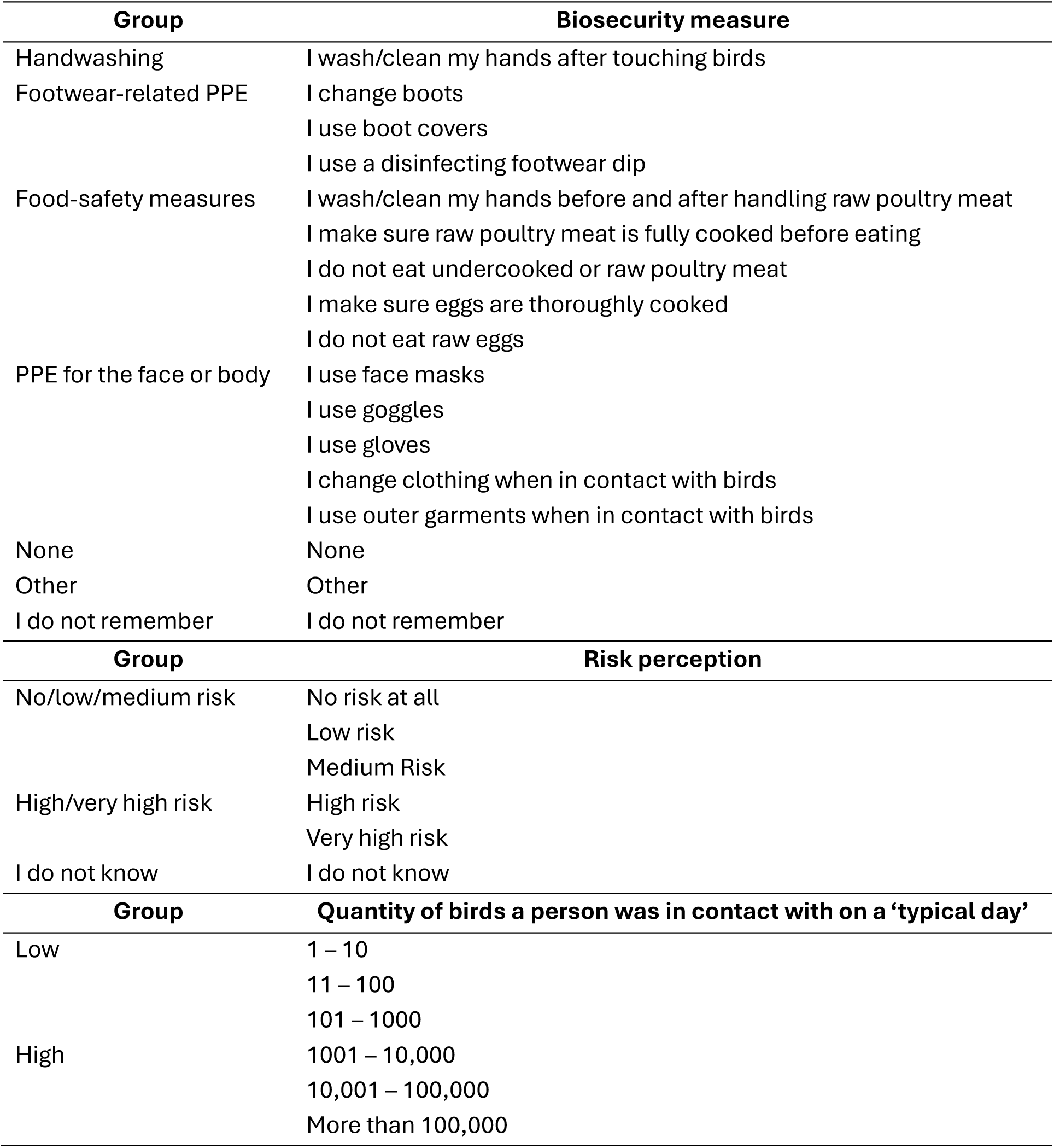
Groupings for biosecurity measures, risk perception and bird contact. Aggregate groups were created for biosecurity measures, risk perception scores and bird contact quantities for inferential analyses.

Descriptive statistics, including frequency counts, proportions and medians were calculated for questions relating to respondent characteristics, bird contact, BMs and risk perception.

Differences in uptake of BMs and risk perception ranks between bird contact groups were tested using the two proportions z-test, and confidence intervals (95%) for a single proportion were also calculated. Chi-squared tests of independence or Fisher’s exact tests (where cell counts were below five) were used to test for an association between the uptake of BMs and the number of birds a person was in contact with (‘bird contact’) alongside perceived risk of AIV (to oneself and birds). Missing responses for BM uptake, bird contact and risk perception were removed from these analyses. The underlying code for data analysis can be found on GitHub (https://github.com/amythomas/aviancontactstudy).

## Results

### Respondent characteristics

A total of 225 respondents completed the questionnaire between 15 May 2024 and 31 July 2024. Demographic characteristics are shown in Table 2. In-person respondents accounted for approximately one third of the sample (28%, 63 of 225) compared to online completion of the questionnaire (72%, 162 of 225). The median age was 48 years, ranging from 19 to 89 years. The largest age category were respondents aged between 50 and 59 (27%, 60 of 225). More than half of responses were from those identifying as male (63%, 142 of 225).

**Table 2.**
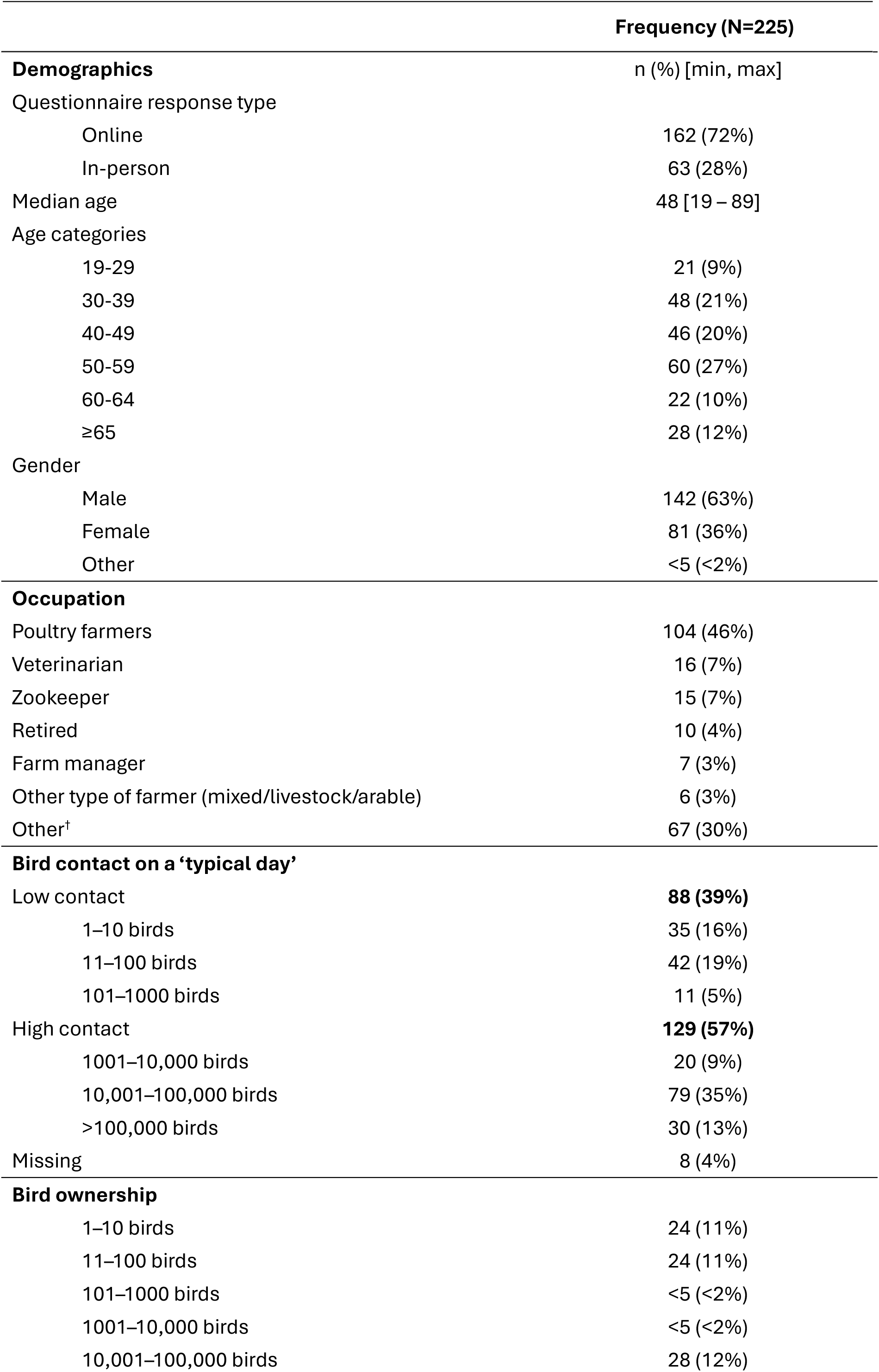
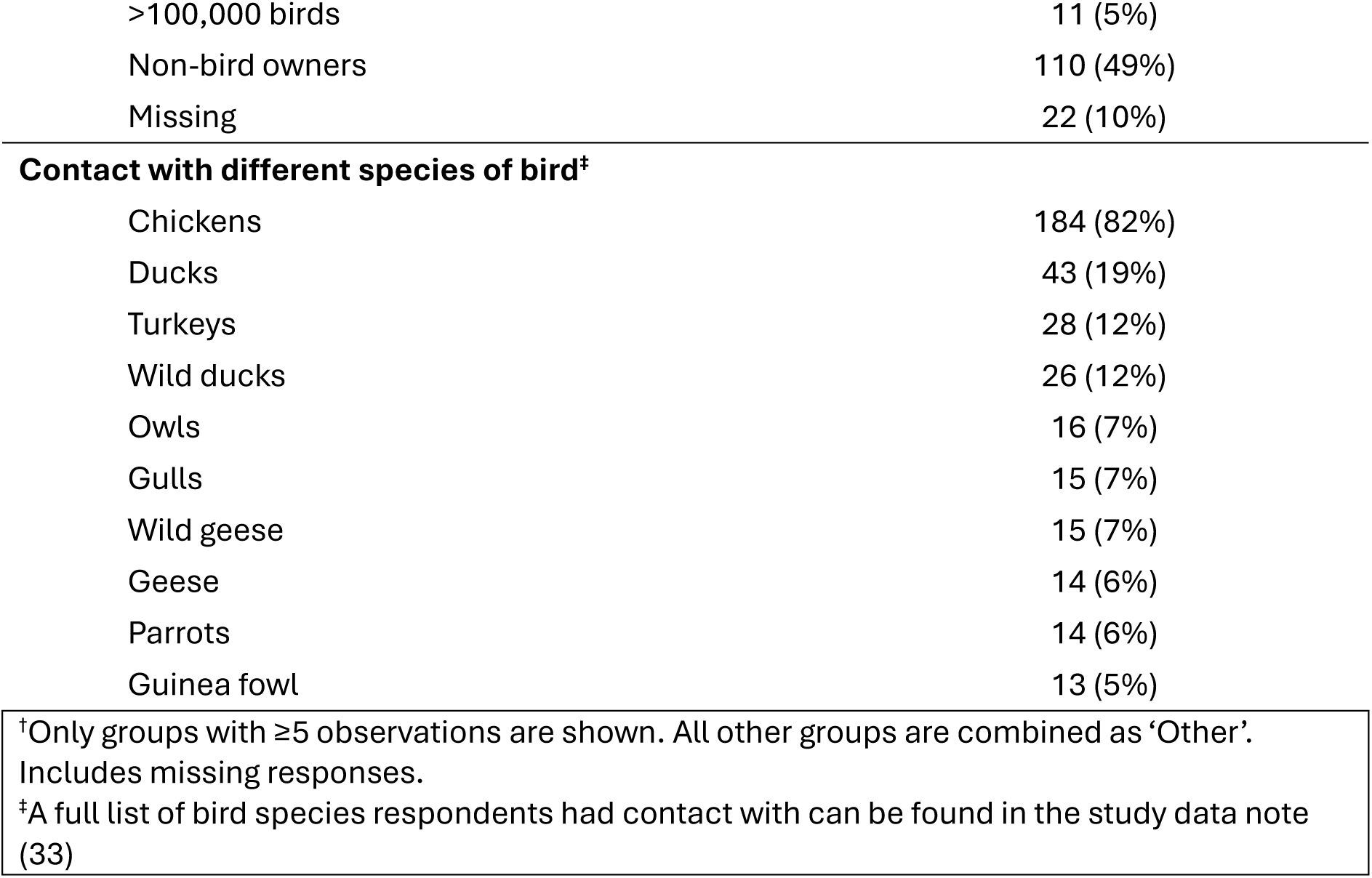
Avian Contact Study respondent characteristics. Note, proportions may not sum to 100% due to rounding.

Respondents provided information on their occupation, bird contact and bird ownership. Almost half were poultry farmers (46%, 104 of 225). Fewer respondents were veterinarians (7%, 16 of 225), zookeepers (7%, 15 of 225), retired (4%, 10 of 225), farm managers (3%, 7 of 225) or another type of farmer (3%, 6 of 225). Other occupations that could not be grouped into major categories accounted for 67 of 225 (30%) respondents, these included roles in business administration, care, hospitality and animal welfare.

Chickens were the single largest species respondents reported contact with (82%, 184 of 225), followed by ducks (both domestic and wild) (19%, 43 of 225), turkeys (12%, 28 of 225), owls (7%, 16 of 225), wild geese (7%, 15 of 225), gulls (7%, 15 of 225), geese (6%, 14 of 225), parrots (6%, 14 of 225) and guinea fowl (5%, 13 of 225). A full list of all the species respondents had contact with are available in the study data note (33).

Over half of respondents reported high contact levels with birds (≥1001 birds/day; 57%; 129 of 225). Low bird contact (≤1000 birds/day) was reported by 39% of all respondents (88 of 225). Respondents were most likely to have daily contact with large flocks of 10,001–100,000 birds (35%; 79 of 225), or presumed hobbyist/back-yard keepers in contact with 1–10 birds (16%; 35 of 225) or 11–100 birds (19%; 42 of 225). There were 8 (4%) missing responses. Most respondents did not own birds (49%; 110 of 225). Of those who did, ownership of birds was evenly distributed across most categories: 11% (24 of 225) owned 1–10 birds, 11% (24 of 225) owned 11–100 birds and 12% (28 of 225) owned 10,001–100,000 birds. There were 22 (10%) missing responses.

### BM measures to limit avian-to-human transmission are influenced by bird contact

Respondents were asked what types of BMs they use to prevent infection with AIV (7 of 225 responses were excluded due to missing BM uptake data). In total, 98% (213 of 218) reported using at least one BM, and 2% (5 of 218) reported using none (Figure 1). The single most reported BM was handwashing after touching birds, used by 89% (196 of 218) of respondents, followed by using disinfecting footwear dips and changing boots, reported by 78% (170 of 218) and 70% (152 of 218), respectively. The proportion of respondents who used at least one BM in each BM group is available in Table S1 of the supplementary material.

**Figure 1.**
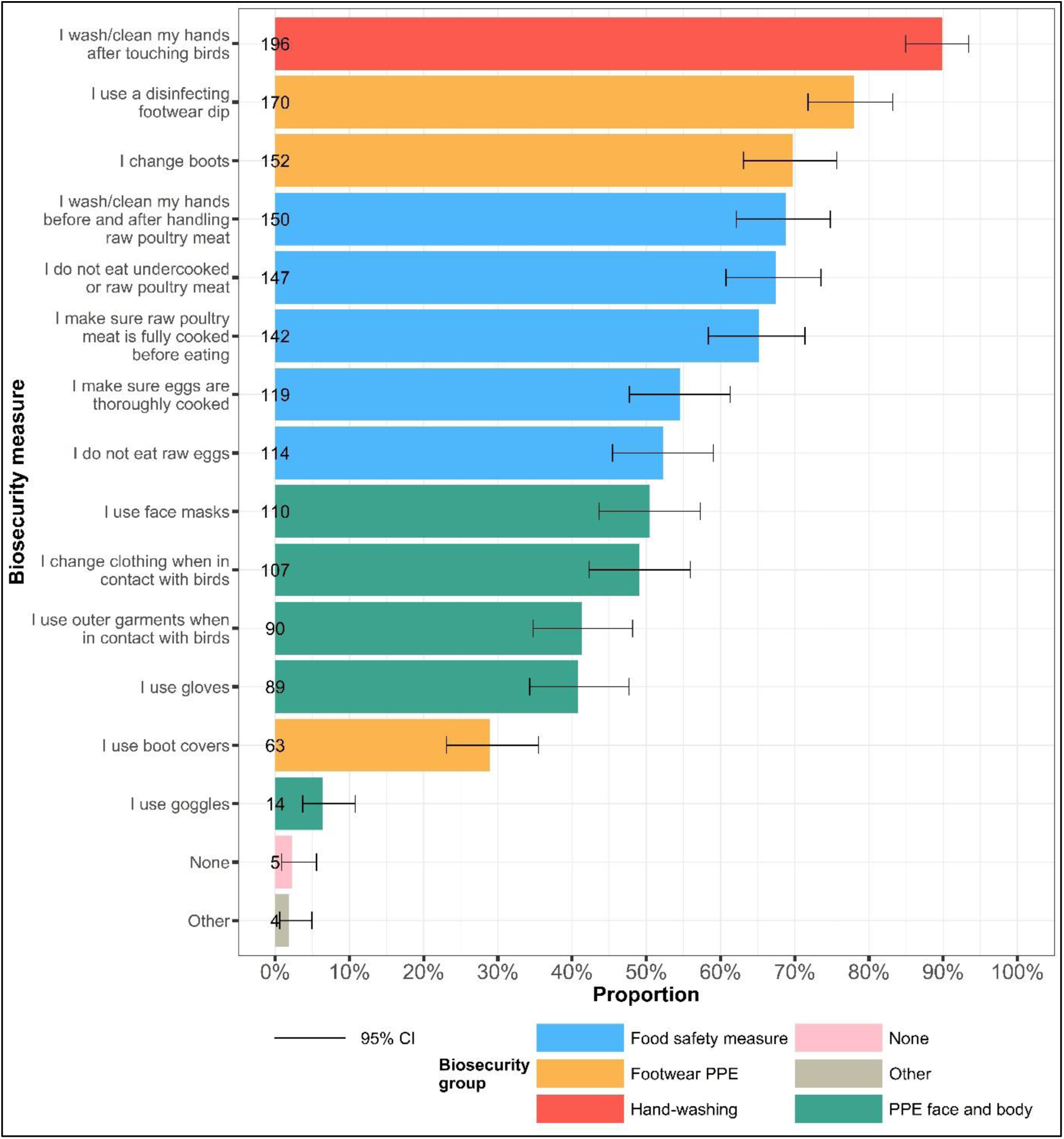
Proportion of participants reporting the use of biosecurity measures to prevent personal infection with avian influenza virus. Colours represent biosecurity measures grouped into similar aggregate categories. Error bars represent 95% confidence intervals for a single proportion.

We hypothesised that the number of birds an individual was in contact with would affect uptake of BMs. A total of 217 respondents provided information on both bird contact and uptake of BMs. There was substantial evidence that a larger proportion of high bird contact respondents (≥1001 birds/day) reported using at least one footwear related PPE measure (z=6.49, p<0.001) or at least one PPE measure for the face or body (z=5.88, p<0.001), compared to low bird contact respondents (Figure 2). There was limited evidence for a difference in the proportion between high and low bird contact respondents reporting the use of at least one food safety measure (z=0.94, p=0.447) and washing their hands after touching birds (z=1.41, p=0.238). Only low bird contact respondents reported using no measures (6%, 5 of 88) (Figure 2).

**Figure 2.**
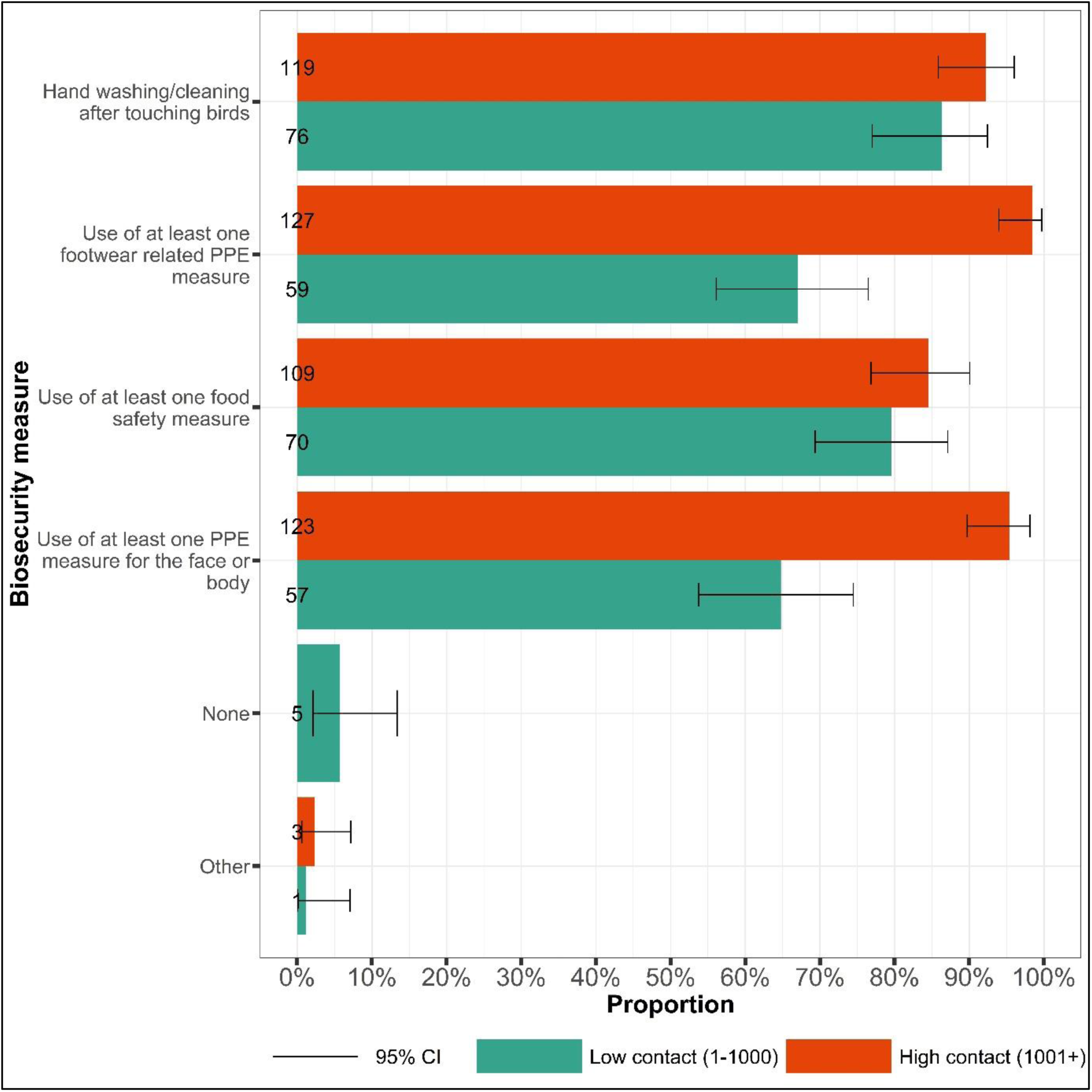
Biosecurity measure uptake by bird contact quantity. Proportion of participants using at least one biosecurity measure as per the aggregate biosecurity categories by low and high bird contact group. Handwashing after touching birds was considered as its own category. Low contact: 1–1000 birds; high contact: ≥1001 birds. Error bars represent 95% confidence intervals for a single proportion.

Uptake of BMs was higher among the high bird contact group than the low bird contact group (Table 3). There was evidence for an association between bird contact levels and the use of at least one PPE measure for the face or body (χ2 (1, n=217) = 32.452, p<0.001), and use of at least one footwear related PPE measure (Df=1, n=217, p<0.001) (Table 3). There was no evidence of association for use of at least one food safety measure (χ2 (1, n=217) = 0.888, p=0.346) or hand washing after touching birds (χ2 (1, n=217) = 1.988, p=0.159). There was strong evidence for the use of no measures (Df=1, n=217, p=0.010). However, frequency counts were small.

**Table 3.**
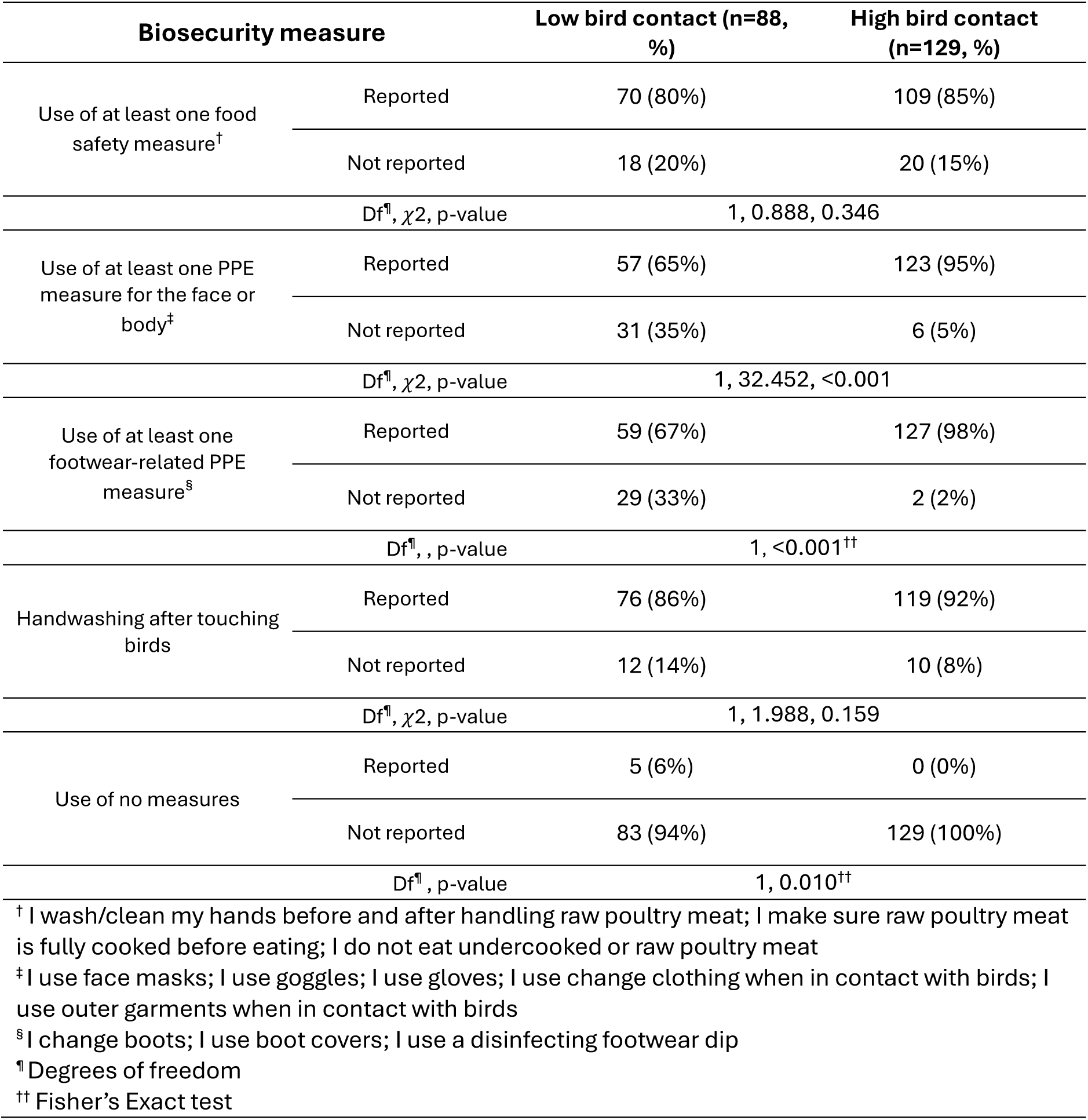
Relationship between biosecurity measures and bird contact. Chi-square test of independence or Fisher’s exact test between biosecurity measures and bird contact level. P- values are rounded to 3 decimal places. Due to rounding, some percentages do not add to 100%.

### Perceptions of avian influenza risk for bird health are higher than human health

We collected respondents’ perceptions of AIV risk to their own health and the health of their birds. After removing missing data and those with unknown bird contact (34 of 225), 116 respondents reported high bird contact, and 75 respondents reported low bird contact (n=191) (Figure 3). Most respondents perceived the risk of AIV to their physical health to be low or medium (80%; 152 of 191). There was moderate evidence that a greater proportion of low bird contact than high bird contact respondents perceived the risk to be low (z=2.38, p=0.017; n-Low=52/75, n-High=59/116). There was no evidence for a difference between bird contact groups in all other risk categories for this question. Risk to the health of respondents’ birds was largely perceived to be medium, high or very high (147 of 191). There was substantial evidence for a greater proportion of low bird contact respondents perceiving the risk to be low compared to high bird contact respondents (z=3.74, p<0.001; n-Low=24/75, n-High=11/116), and for increased high bird contact respondents perceiving the risk to be very high compared to low bird contact respondents (z=3.53, p<0.001; n-Low=8/75, n-High=40/116).

**Figure 3.**
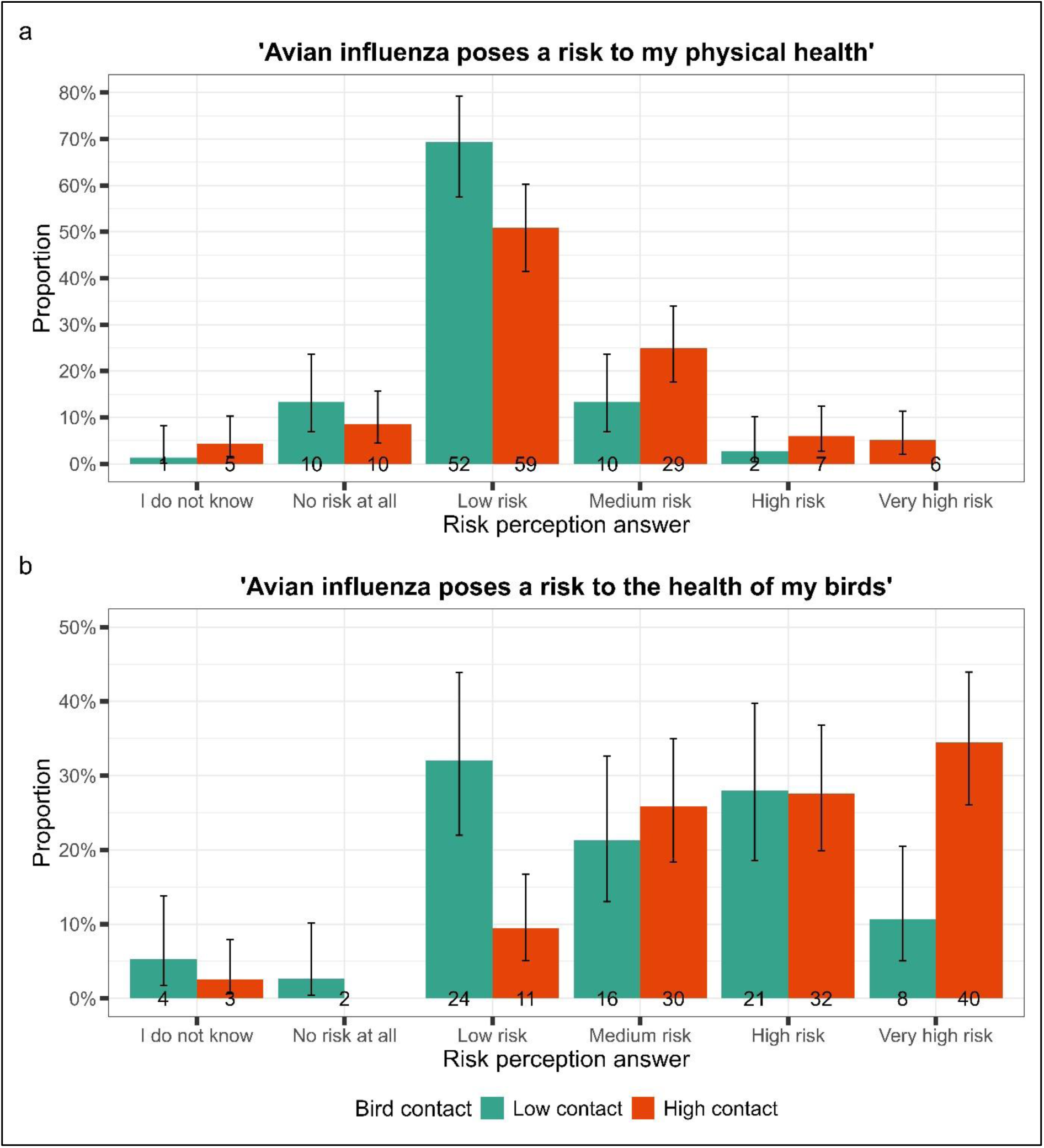
Risk perception of avian influenza by bird contact. Proportion self-reporting agreement with risk statements by bird contact group (low and high) for a) risk to human health and b) risk to the health of their birds. Low contact: 1 to 1000 birds; high contact: ≥1001 birds. Error bars represent 95% confidence intervals for a single proportion.

Further missing responses in the physical health (n=5) and bird health (n=6) categories were removed, resulting in 186 and 185 respondents respectively. Respondents who considered AIV risk as high to bird health were more likely to use at least one footwear related PPE measure than those who perceived the risk as low (χ2 (1, n=185)= 9.171, p=0.002). For all other biosecurity groups, there was no evidence of an association with perceived risk to bird health or perceived risk to physical health (Table 4).

**Table 4:**
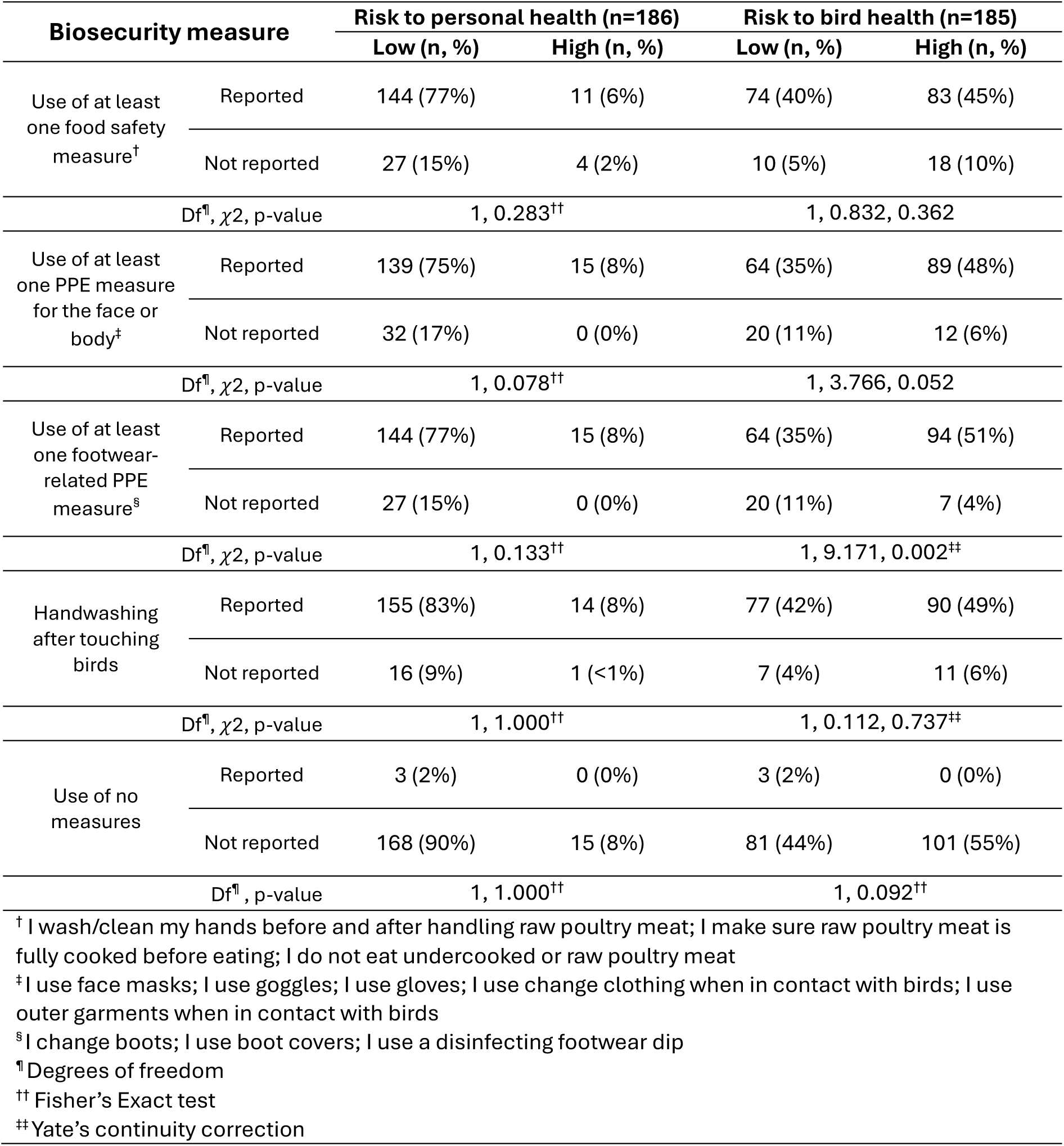
Relationship between biosecurity measure and perceived risk of avian influenza virus to bird and personal health. Chi-squared or Fisher’s exact tests of association between the use of biosecurity measures and perceived risk of avian influenza virus to personal and bird health. Biosecurity measures were grouped into themes where appropriate. Risk groups were aggregated into low (no, low and medium) and high (high and very high) risk. P-values are rounded to 3 decimal places. Due to rounding, some percentages do not add to 100%.

## Discussion

We determined biosecurity practices and risk perceptions of zoonotic avian influenza virus (AIV) among individuals in contact with kept and wild birds in the UK. Our analyses identified that handwashing was the most common biosecurity measure (BM) reported by respondents to prevent infection with AIV, followed by use of disinfecting footwear dips and changing boots. Almost all respondents reported using at least one BM, and uptake was higher for respondents in daily contact with high numbers of birds. The perceived risk of AIV to respondents’ own health was low, regardless of the number of birds a respondent had daily contact with. Whereas perceived risk of AIV to bird health was high. Our findings suggest that BM uptake in people in contact with birds is likely influenced by the perceived risk to avian health, but not human health, and is further impacted by bird contact/farming scale.

The reported high uptake of footwear-related PPE measures is consistent with a recent qualitative study of UK poultry farmers (21), strengthening our understanding of these being common practices among the poultry industry. In contrast to a European study on poultry farms, we found hand washing after touching birds was the most frequently reported BM (24). This could reflect differences in questionnaire framing, where questions intended to elicit answers for preventing avian-to-human transmission led to respondents considering personal hygiene activities with improved response efficacy, as opposed to avian-centred BMs (26). This suggests that public health campaigns framed around avian-to-avian transmission risk may encourage the uptake of different BMs compared to those centred on avian-to-human transmission risk. In turn, this could affect the ability to control onward transmission of AIV following spillover to people and highlights the importance of appropriate risk framing.

Overall, it is reassuring that 98% of respondents reported using at least one BM. This could be indicative of prevalent risk assessment practices among our sample, or a social desirability bias induced by face-to-face data collection when asked about actions to prevent avian-to-human disease transmission. Interestingly, we found that 83% of respondents reported use of at least one food safety measure. Understanding implementation of food safety in this cohort in the absence of targeted public health messaging specifically for AIV in the UK provides a baseline of practice. This is important given AIV transmission among dairy cattle and contaminated dairy products (15). Across all BMs, uptake differed by bird contact and show that daily contact with a high number of birds was associated with greater BM uptake to prevent zoonotic AIV transmission. As a proxy for farming operation size, this is similar to other studies where larger farm sizes report higher BM uptake for the prevention of avian-to-avian transmission (23,28). Although small flock systems present a lower risk of AIV outbreaks (22), lower BM uptake may put individuals in contact with birds at a greater risk of exposure. Identifying cohorts with low BM uptake, like those in contact with small flocks, is useful to optimise targeted preventive public health programmes for zoonotic AIV and improve uptake of multiple BMs. The perceived risk of AIV to human health was not associated with the use of any BM measure, whereas risk to bird health was associated with the uptake of at least one footwear-related PPE measure. Using protection motivation theory (26), this may be explained by participants’ perceived susceptibility and severity of AIV to human health, demonstrated by risk rankings being low and medium, which has been observed in other demographics (31). This is in line with other findings that perceived human susceptibility to AIV is low, possibly affecting uptake of certain protective behaviours (39). This reinforces the importance of how the public health narrative of zoonotic AIV is framed when increased BM uptake is a desired outcome. Moreover, limited evidence for sustained human-to-human transmission may result in little motivation to use BMs (13).

## Strengths and limitations

This study compliments existing knowledge regarding biosecurity usage to prevent avian and human infection among bird keepers in Europe and South East Asia (22,31), and the changing transmission dynamics in North America and Europe over the last four years (3,12,13). Our questionnaire was co-developed with poultry farmers, veterinary public health agencies and stakeholders (33). This approach ensured relevancy of questions, accessibility of language and provided timely insight to the behaviours of groups potentially at risk of AIV exposure. Moreover, engagement with the commercial poultry sector has enabled us to generate evidence of their biosecurity practices, useful for informing ongoing policy developments that aim to curb high-risk occupational exposures to AIV in commercial poultry settings.

Although individuals in contact with any birds (wild and/or domestic) could participate, there was an overrepresentation of commercial poultry farmers in the sample. Accordingly, chickens were the most frequently reported species that respondents had contact with. This is likely a feature of launching the questionnaire at a national Pig & Poultry Fair (33), limiting the generalisability of our findings to small-flock holders, like those studied in the UK Flockdown survey (22) and wild bird keepers. Some covariates, such as farm assurance schemes and previous AIV outbreaks were not measured, preventing some statistical tests from being robustly performed. Future iterations of the study will aim to record these and ensure that questions are time-bound to enable the assessment of consistent biosecurity uptake. This will improve our understanding of how live outbreaks and wider public health contexts influence behaviour.

## Conclusions

Our findings suggest that use of BMs by people who have contact with birds in the UK are influenced by the number of birds a person is in contact with (as a proxy for farming operation size) and the perceived risk to bird health (high), but not perceived risk to human health (low). Further work should be undertaken to identify the characteristics which lead to low BM uptake among different cohorts in contact with different bird species to help inform zoonotic AIV public health messaging and campaigns. Moreover, a more extensive investigation into the barriers and facilitators of BM uptake is vital for improving our understanding of nuanced behaviour in the context of zoonotic influenza spillover.

## Data availability

Repository data.bris: The Avian Contact Study: questionnaire data 15 May – 31 July 2024. Data are openly available at the University of Bristol Research Data Repository data.bris, at https://doi.org/10.5523/bris.3nmqsrbv5ruom2abn0ql6e8yh2.

This project contains the following underlying data:

- - Data file 1. (Raw underlying questionnaire data – csv file)
- - Data file 2. (Raw underlying questionnaire data – .RDS file)
- - Data file 2. (Associated data dictionary – csv file)
- - Data file 3. (Code for importing underlying data in csv format into R for setting up labelled data – .r file)
- - Data file 4. (Blank consent form and participant information sheet – pdf file)

Data are available under the terms of National Archives’ Non-Commercial Government Licence for public sector information.

## Software availability

Source code available from: https://github.com/amythomas/aviancontactstudy.

## Conflicts of interest

LES, SG, SM, JT and RP are employees of the UK Health Security Agency. LES receives consultancy fees from the Sanofi group of companies and other life sciences companies. PM is an employee of the Animal Plant and Health Agency. The views expressed are those of the authors and not necessarily those of the UKHSA or the Department of Health and Social Care.

## Funding statement

Funding for the Avian Contact Study was awarded by PolicyBristol from the Research England QR Policy Support Fund (QR PSF) 2022-24 for investigating ‘Zoonotic spillover of avian influenza’. AT is funded by the Wellcome Trust, Early Career Award [227041/Z/23/Z]. EBP acknowledges support from the National Institute for Health Research Health Protection Research Unit (NIHR HPRU) in Evaluation and Behavioural Science at the University of Bristol (NIHR207385).

## Acknowledgements

We thank all participants who completed the questionnaire and those who contributed to the development of the study.

## Supplementary materials

1. aiv_biosecurity_supplementary.docx - document containing Figure S1 (frequency counts of different biosecurity measures in each biosecurity group), Figure S2 (frequency counts of risk perception categories for risk of avian influenza to respondents’ health and the health of their birds) and Table S1 (Frequency of respondents who used at least one biosecurity measure from each biosecurity group)

## Notes

### Author Declarations

Ethical approval for the study was obtained from the University of Bristol, Faculty Research Ethics Committee, approval number 17048 on 16 January 2024. Informed written consent (using e-consent hosted on Research Electronic Data CAPture tools, REDCap) for the use of data collected via the questionnaire was obtained from respondents.

### Summary of Updates

Extra detail on cleaning and pre-processing given in the methods section; inclusion of the gender split of respondents and the species of birds they had contact with in the results section; comments on food safety measures and the uptake of at least one biosecurity measure in the discussion; imrpoved clarity of Figures in the main text and supplemental material; correction of author affiliations; and an additional table in the supplemental material.

