## Supplemental Figures S1, S2 and Table S1 for "Biosecurity uptake and perceived risk of avian influenza among people in contact with birds"

5: UK NIHR Health Protection Research Unit in Behavioural Science and Evaluation

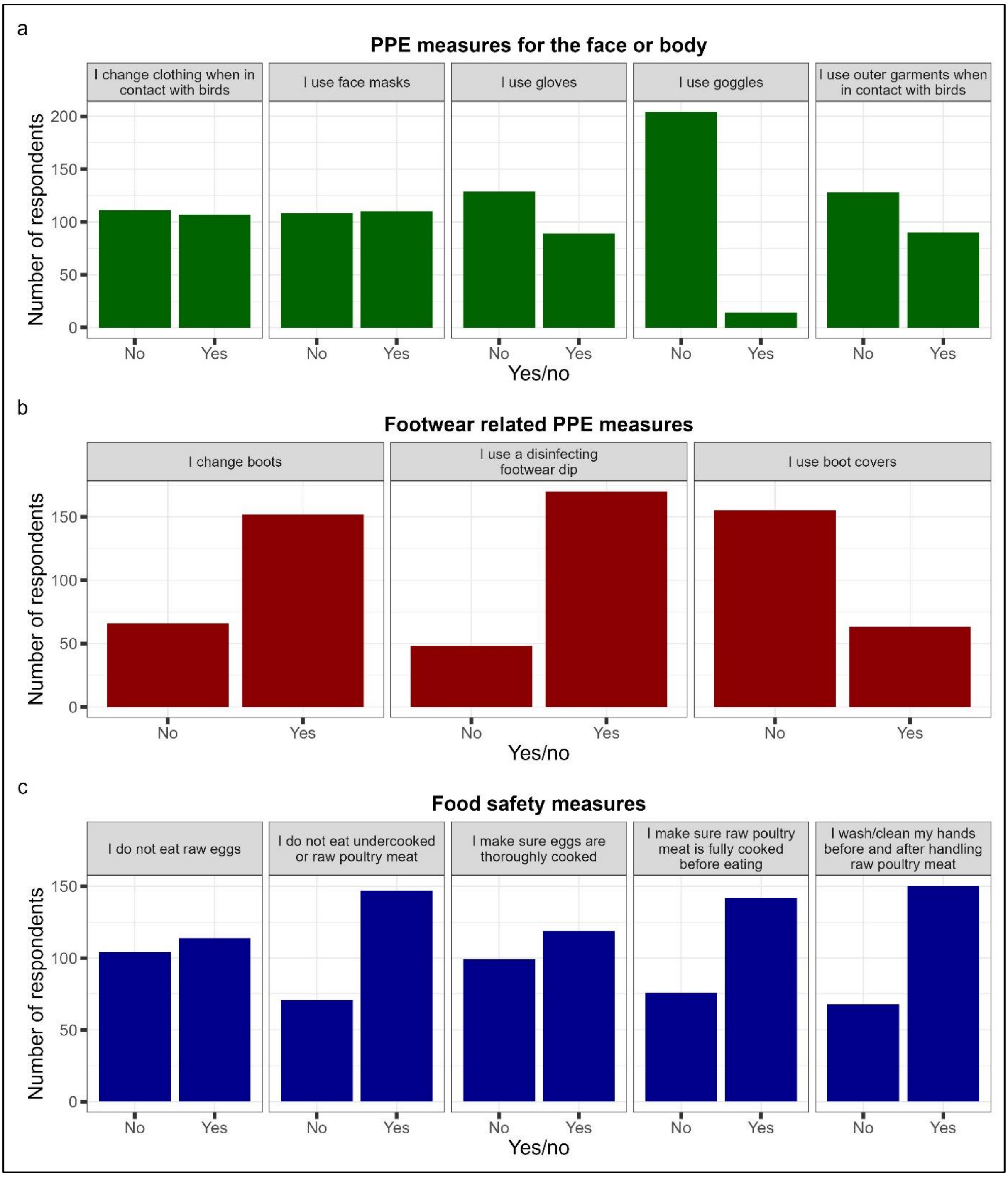


**Figure S1.** Frequency counts of different biosecurity measures in each biosecurity group. (a) Use of at least one PPE measure for the face or body, (b) use of at least one footwear related PPE measure and (c) use of at least one food safety measure. Use of biosecurity measures were recorded using radio buttons that respondents "checked" for yes. We assumed responses with all measures "unchecked" indicated a skipped question and were therefore coded as missing.

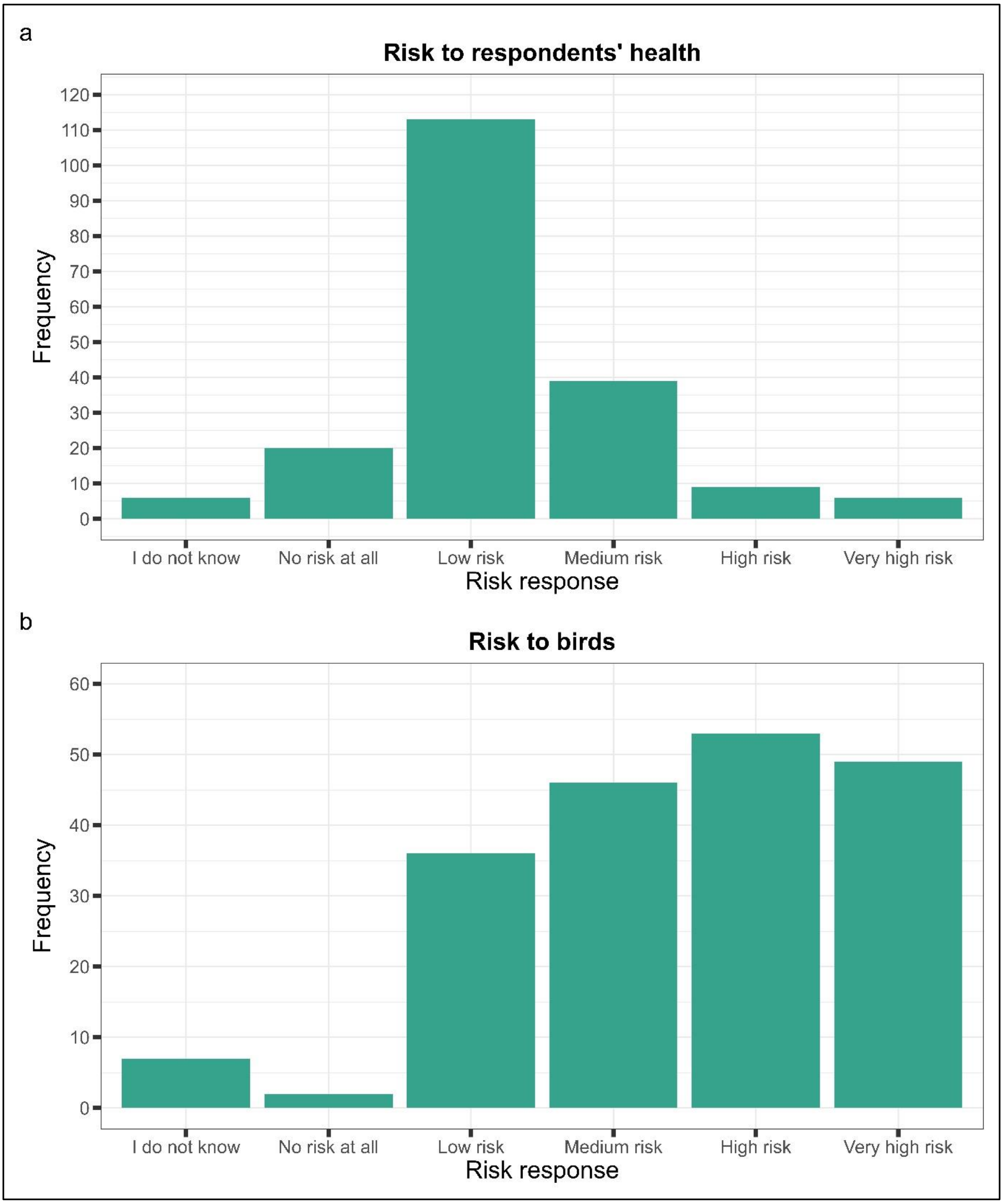


**Figure S2.** Frequency counts of risk perception categories for risk of avian influenza to (a) respondents' health and (b) risk to the health of their birds.

**Table S1.** Frequency of respondents who used at least one biosecurity measure from each biosecurity group.

| Biosecurity measure group | Number of respondents using at least one measure in the group (n=217): |
|---|---|
| PPE measures for the face or body | 181 (83.4%) |
| Footwear related measures | 187 (86.2%) |
| Food safety measures | 180 (82.9%) |
